# Whole-exome-based preconception carrier screening in Uzbekistan with targeted SMA, FMR1, and DMD assays: the first reported clinical program

**DOI:** 10.64898/2026.06.02.26354713

**Authors:** Andrei Kullyev, Svetlana Avdeichik, Alexandra Akimenkova, Andrey Kartuesov, Olga Kardymon, Yaron Goikhman

## Abstract

**Purpose:** Published clinical outcome data on preconception carrier screening (PCS) in Central Asia are limited. We report the first clinical implementation study from Uzbekistan of a whole-exome sequencing (WES)-based multi-platform PCS program combining exome sequencing with targeted SMA, FMR1, and DMD assays.

**Methods:** We retrospectively analyzed anonymized data from 65 individuals (19 couples, 27 singletons) screened at IMC Genomics, Tashkent, between January 2024 and May 2026. WES covering the protein-coding regions of approximately 20,000 genes was followed by exome-wide bioinformatics filtering and clinical geneticist interpretation. Partly overlapping cohorts underwent SMA carrier screening (n=179), FMR1 CGG-repeat analysis in females (n=155), and DMD deletion/duplication testing in preconception females (n=29). Variants were classified by ACMG/AMP criteria against gnomAD v4.1.

**Results:** Sixty-one of 65 WES-screened individuals (93.8%; 95% CI 85.2–97.6%) carried at least one reportable variant (152 instances across 126 genes). Four of 19 couples (21.1%; 95% CI 8.5–43.3%) were concordant for pathogenic or likely pathogenic variants in the same autosomal recessive gene; two were referred for preimplantation genetic testing for monogenic disease. SMA screening identified four carriers, including two 2+0 silent carriers; FMR1 analysis identified one intermediate allele; DMD MLPA identified no exonic rearrangements.

**Conclusion:** This first reported WES-based multi-platform PCS program in Uzbekistan was feasible and clinically informative, identifying actionable couple-level reproductive risks and supporting structured implementation of reproductive genetic screening in Central Asia.

## Introduction

Preconception carrier screening (PCS) enables couples to identify their carrier status for serious autosomal recessive and X-linked conditions before or during early pregnancy, providing the opportunity to make informed reproductive decisions and, where indicated, access preimplantation genetic testing for monogenic disease (PGT-M) or prenatal diagnosis. Expanded PCS programs using whole-exome sequencing (WES) or large gene panels have been implemented across North America, Western Europe, Israel, and Australia, demonstrating carrier detection rates consistently exceeding 90% for expanded panels (Tong et al. 2022) and concordant at-risk couple identification rates of approximately 2–4% in outbred populations (Capalbo et al. 2021). While most expanded PCS programs rely on targeted gene panels or medical exomes, a comprehensive exome-based filtering strategy has been successfully applied in both consanguineous and non-consanguineous couples (Sallevelt et al. 2017).

Consanguineous marriage is globally associated with elevated risk of autosomal recessive disease; approximately 10% of all marriages worldwide are contracted between second cousins or closer (Bittles and Black 2010). In Muslim-majority populations spanning the Middle East and Central Asia, rates of consanguineous marriage can reach 25–30% of all unions, driven by long-standing socio-cultural tradition (Tadmouri et al. 2009). In Uzbekistan specifically, consanguineous marriage rates as high as 71.2% have been documented in a rural population of Samarkand province (Paradeeva and Revazov 1987). Consistent with this recessive-disease burden, a survey of 671 children attending schools for the blind in Uzbekistan found consanguinity in 33.2% of blind and severely visually impaired cases and hereditary disease (retinal dystrophies and familial cataract) in 54.5% of blindness cases, with retinal dystrophies alone accounting for 24% of cases; the high proportion of retinal dystrophies was attributed to the common practice of consanguineous marriage (Rogers et al. 1999).

Beyond the amplification of common variants, the Uzbek population remains largely uncharacterized with respect to its pathogenic variant landscape. Published population genomic data for Central Asian populations are sparse; gnomAD v4.1 contains limited South Asian and East Asian reference data but no dedicated Central Asian cohort. This means that pathogenic variant frequencies relevant to Uzbek patients cannot be reliably inferred from existing databases, and population-specific founder alleles are unlikely to be identified without targeted clinical characterization.

To our knowledge, this is the first clinical implementation study from Uzbekistan of a whole-exome sequencing (WES)-based multi-platform PCS program. Full WES covering the protein-coding regions of approximately 20,000 genes was used as the molecular backbone, with bioinformatics filtering applied to the entire exome followed by clinical geneticist interpretation. The program was implemented at IMC Genomics, Israeli Medical Center Department of Human Genetics, Tashkent, Uzbekistan. It was supplemented by targeted assays for conditions where WES has insufficient analytical sensitivity: SMA carrier screening (CarrierMax™ SMN1/SMN2 multiplex PCR/capillary electrophoresis), FMR1 CGG-repeat analysis for Fragile X syndrome, and DMD exonic deletion/duplication screening (SALSA MLPA, MRC Holland).

## Materials and Methods

### Study Design and Ethical Framework

This is a retrospective analysis of anonymized clinical data generated through routine preconception carrier screening at IMC Genomics, Tashkent, Uzbekistan, between January 2024 and May 2026. The study was conducted in accordance with the Declaration of Helsinki. Ethics approval details are provided in the Statements and Declarations section.

### Study Participants

The WES cohort comprised 65 individuals who underwent expanded carrier screening across 14 consecutive sequencing runs. Participants presented either as couples or as singletons following pre-test genetic counseling, at which point they selected the testing components they wished to undergo (menu-based model). Nineteen couples (38 individuals) and 27 singletons were included. Two laboratory staff members who self-tested through the same pipeline were excluded from analysis to prevent ascertainment bias. Additional, larger cohorts were independently screened for SMA, Fragile X (FXS), and DMD using dedicated targeted assays; these cohorts overlap with but extend beyond the WES cohort and are reported with separate denominators.

### WES-Based Carrier Screening

Genomic DNA was extracted from peripheral blood leucocytes using the QIAamp DNA Mini Kit (Qiagen). Exome libraries were prepared using the Illumina Exome enrichment kit and sequenced on an Illumina NextSeq 2000 platform (2×150 bp paired-end). Mean coverage depth was approximately 160× (range 153–173× across runs), with ≥93% of targeted bases at ≥30×. Reads were aligned to GRCh38/hg38. Variant calling and annotation were performed using Emedgene (Illumina), a validated clinical interpretation platform for next-generation sequencing analysis. The bioinformatics filtering strategy applied to the entire exome was adapted from previously published approaches for preconception carrier screening (Sallevelt et al. 2017). Candidate variants were filtered across the entire exome at an allele frequency threshold of ≤1% in gnomAD v4.1 (Karczewski et al. 2020) for autosomal recessive conditions (≤0.1% for severe early-onset conditions). Candidate variants meeting these filtering criteria were then reviewed and interpreted by clinical geneticists. Only pathogenic (P) and likely pathogenic (LP) variants in genes associated with serious autosomal recessive or X-linked recessive disorders were reported. Variant classification followed ACMG/AMP Standards and Guidelines (Richards et al. 2015), referenced against gnomAD v4.1 and ClinVar (Landrum et al. 2016). VUS were excluded from carrier frequency calculations. Secondary findings as defined by the ACMG SF v3.3 list (Lee et al. 2025) were reported separately in Supplementary Table 2. For X-linked genes, hemizygous variants in male participants were interpreted as indicating an affected genotype rather than carrier status. One complex cis allele (GNRHR c.[317A>G;785G>A]) was counted as a single carrier event, and the BTD c.1270G>C (p.Asp424His) pseudodeficiency allele was excluded from carrier frequency calculations. All variants met pre-established quality thresholds for clinical reporting. The IMC Genomics laboratory operates under an institutional quality management framework consistent with ISO 15189 principles. This study was reported in accordance with STROBE guidelines for observational studies.

### SMA Carrier Screening

SMA carrier status was assessed using the CarrierMax™ SMN1/SMN2 Reagent Kit (Thermo Fisher Scientific/Applied Biosystems), which employs multiplex PCR amplification of genomic DNA with fluorescently labelled primers followed by capillary electrophoresis fragment analysis on a SeqStudio Genetic Analyzer. This assay quantifies SMN1 and SMN2 exon 7 copy number and simultaneously interrogates two intragenic SNP variants associated with the silent carrier (2+0) haplotype, including g.27134T>G (rs143838139), enabling discrimination of the 2+0 cis (silent carrier) configuration from a normal 1+1 trans configuration. Regular carriers (1+0 genotype) were distinguished from silent carriers (2+0 genotype, identifiable only through the combined copy-number and SNP-haplotype analysis). Amplicons were sized and quantified using GeneMapper Software in conjunction with CarrierMAX Software. SMA screening was offered independently of WES; the SMA cohort is reported with its own denominator.

### Fragile X Syndrome Screening

FMR1 CGG repeat analysis was performed using the CarrierMax™ FMR1 Reagent Kit (Thermo Fisher Scientific/Applied Biosystems) by PCR, with amplicons resolved by capillary electrophoresis on a SeqStudio Genetic Analyzer. Per institutional protocol, FMR1 analysis was performed in female participants only. CGG alleles were classified as: normal (<45 repeats), intermediate (45–54), premutation (55–200), or full mutation (>200), consistent with ACMG guidelines.

### DMD Carrier Screening

DMD exonic deletion/duplication carrier status was assessed in preconception females by MLPA using the SALSA MLPA Probemix P034-B2 DMD-1 and P035-B1 DMD-2 (MRC Holland) probe sets in combination, covering all 79 exons of the DMD Dp427m muscle transcript, exon 1 of Dp427c, and reference probes (97 probes total). Amplicons were resolved on a SeqStudio Genetic Analyzer. DMD MLPA detects large intragenic deletions and duplications (~65–70% of pathogenic DMD variants); point mutations and small indels were simultaneously assessed through the WES component for participants who underwent both assays. Diagnostic cases (amniocentesis, maternal testing of a known proband) were excluded from the preconception carrier frequency denominator.

### Couple-Level Concordance Analysis

For couples where both partners completed WES-based carrier screening, a joint concordance analysis was performed. A couple was classified as concordant at-risk if both partners carried P or LP heterozygous variants in the same gene associated with the same AR condition (25% per-pregnancy recurrence risk). Concordant couples were referred for post-test genetic counseling and, where indicated, for PGT-M.

### Statistical Analysis

Carrier frequencies were calculated separately for each platform using the total number of individuals completing that assay as the denominator. Binomial 95% confidence intervals were calculated using the Wilson score method. The couple-level concordance rate was calculated as the proportion of concordant couples among all screened couples (4/19). No formal multiple testing correction was applied; all comparisons are descriptive and hypothesis-generating. All computations were performed in Python 3.12 (scipy.stats module).

## Results

### Study Population

Participant flow and assay overlap are summarized in Figure 1. Between January 2024 and May 2026, 65 individuals underwent WES-based carrier screening across 14 sequencing runs at IMC Genomics. The cohort comprised 19 couples (38 individuals) and 27 singletons. Of the 65 participants, 36 were female and 29 were male. Sex distribution is relevant to the targeted-assay denominators, as FMR1 and DMD screening were offered to female participants only. Participants represented multiple ethnic backgrounds including Uzbek, Russian, Korean, and other nationalities, consistent with the diverse urban population of a private clinical genetics center in Tashkent. Fifty-eight of the 65 WES-screened participants also underwent SMA carrier screening, 31 of the 36 female WES participants also underwent FMR1 CGG-repeat analysis, and 15 of the 36 female WES participants also underwent DMD deletion/duplication MLPA. The SMA (n=179), FXS (n=155), and DMD (n=29) assay cohorts are larger than the WES cohort because these targeted assays are substantially less expensive than WES and were offered independently to a broader set of patients attending the center, including many who did not pursue exome sequencing. Each is reported with its own denominator.

**Figure 1.**
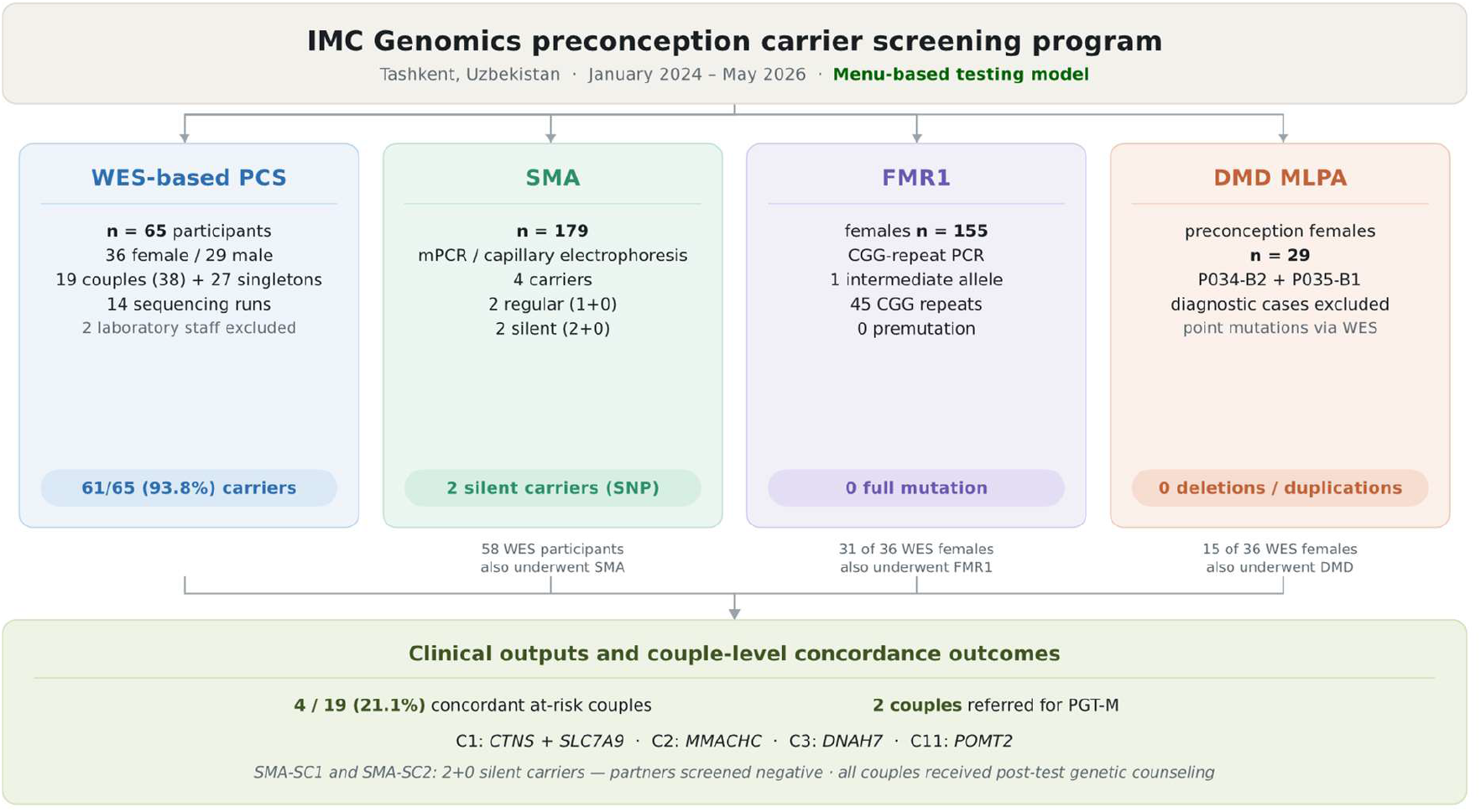
Participant flow and assay overlap in the IMC Genomics preconception carrier screening program. The WES-based PCS cohort comprised 65 individuals screened across 14 sequencing runs, including 19 couples (38 individuals) and 27 singletons; two laboratory staff members who self-tested through the same pipeline were excluded from analysis. Targeted assays were offered independently under a menu-based model with separate denominators: SMA carrier screening, n=179; FMR1 CGG-repeat analysis in females, n=155; and DMD deletion/duplication MLPA in preconception females, n=29. Fifty-eight of the 65 WES-screened participants also underwent SMA testing, 31 of the 36 female WES participants also underwent FMR1 analysis, and 15 of the 36 female WES participants also underwent DMD MLPA. Diagnostic DMD cases were excluded from the preconception denominator. The bottom panel summarizes couple-level concordance outcomes from the WES cohort. WES, whole-exome sequencing; PCS, preconception carrier screening; SMA, spinal muscular atrophy; FMR1, fragile X messenger ribonucleoprotein 1; DMD, Duchenne muscular dystrophy; MLPA, multiplex ligation-dependent probe amplification; PGT-M, preimplantation genetic testing for monogenic disease; SNP, single-nucleotide polymorphism.

### WES-Based Carrier Screening

#### Overall yield

Sixty-one of 65 individuals (93.8%; 95% CI 85.2–97.6%) carried at least one P or LP variant in an AR or X-linked recessive disease gene. Four individuals (6.2%; 95% CI 2.4–14.8%) had no reportable findings. All four had adequate sequencing quality (mean depth 153–173×, ≥93% of targets at ≥30×), confirming true negative results. A total of 152 P/LP variant instances were identified across 126 unique disease genes. The mean number of reportable variants per carrier was 2.5 (range 1–6).

#### Most frequently implicated genes

The gene most commonly affected was CFTR (cystic fibrosis; MIM 219700), with P or LP variants detected in 5 participants (7.7%) across five distinct variants: c.3935A>G (p.Asp1312Gly), c.413_415dup (p.Leu138dup), c.3154T>G (p.Phe1052Val), c.2856G>C (p.Met952Ile), and c.1367T>C (p.Val456Ala). GJB2 (connexin 26; deafness DFNB1; MIM 220290) was the second most frequently affected gene with 4 carriers (6.2%) carrying four distinct variants: c.35del (p.Gly12ValfsTer2, the globally prevalent 35delG allele; frequency 0.70% in gnomAD v4.1), c.−23+1G>A (P; ClinVar ID 17029), c.101T>C (p.Met34Thr; LP), and c.95G>A (p.Arg32His; LP). HFE (hereditary haemochromatosis; MIM 235200) was identified in one participant of Russian ethnicity, carrying the p.Cys282Tyr founder allele (gnomAD v4.1 non-Finnish European frequency 5.7%; ClinVar rs1800562).

#### Rare or gnomAD-absent variants of regional interest

Several rare or gnomAD-absent variants were observed. USH2A c.13339A>G (p.Met4447Val; absent from or extremely rare in gnomAD v4.1) was detected in two unrelated participants across different sequencing runs. CTNS c.225+1G>A (absent from gnomAD v4.1) was found in both members of Couple 1, representing a shared couple-level finding. PLEKHG2 c.610C>T (p.Arg204Trp; absent from gnomAD v4.1) and LIPH c.742C>A (p.Leu248Met; a characterized East Asian founder allele, Shimomura et al. 2009) were each identified in one participant. These observations are hypothesis-generating and require confirmation in larger ancestry-stratified Central Asian cohorts with haplotype analysis before population-enriched or founder status can be assigned.

#### X-linked finding

One male participant was identified as hemizygous for G6PD c.1003G>A (p.Ala335Thr; ClinVar ID 10363; MIM 300908), consistent with G6PD deficiency. G6PD p.Ala335Thr (ClinVar 10363) is classified as Pathogenic for G6PD deficiency; WHO Class II (G6PD Chatham variant), with <10% residual enzyme activity and associated risk of acute hemolytic episodes on oxidative stress exposure. This finding is reported separately and excluded from AR carrier frequency calculations. His female partner carried no G6PD pathogenic variant by WES. This participant is counted as a carrier on the basis of four heterozygous LP variants in AR disease genes (DNAH7, OAT, CBLIF, RAG1).

#### Secondary findings

ACMG SF v3.3 secondary findings were identified in five participants (Supplementary Table 2): TTR c.424G>A (p.Val142Ile; gnomAD v4.1 African frequency >1.7%); PMS2 c.251-1G>T (Lynch syndrome; NM_000535.7; variant call confirmed against PMS2 reference sequence to exclude PMS2CL pseudogene cross-mapping); SOD1 c.272A>C (ALS type 1); CHEK2 c.470T>C (p.Ile157Thr); and MYH7 c.968T>C (p.Ile323Thr). All were disclosed in accordance with institutional protocol. MLH3 c.1755del, identified in one additional participant, is not currently on the ACMG SF v3.3 list but was disclosed clinically given its potential mismatch repair association; its Lynch syndrome relationship is less robustly established than that of MLH1, MSH2, MSH6, or PMS2.

**Table 1:**
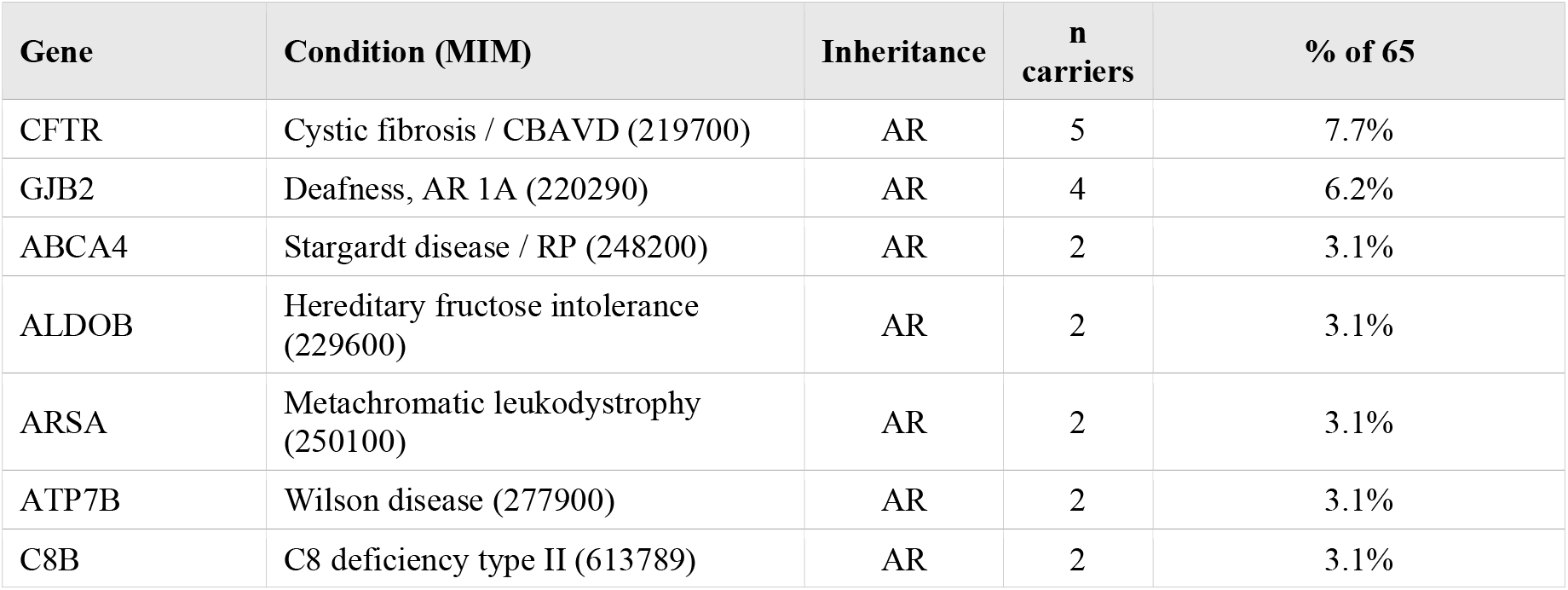
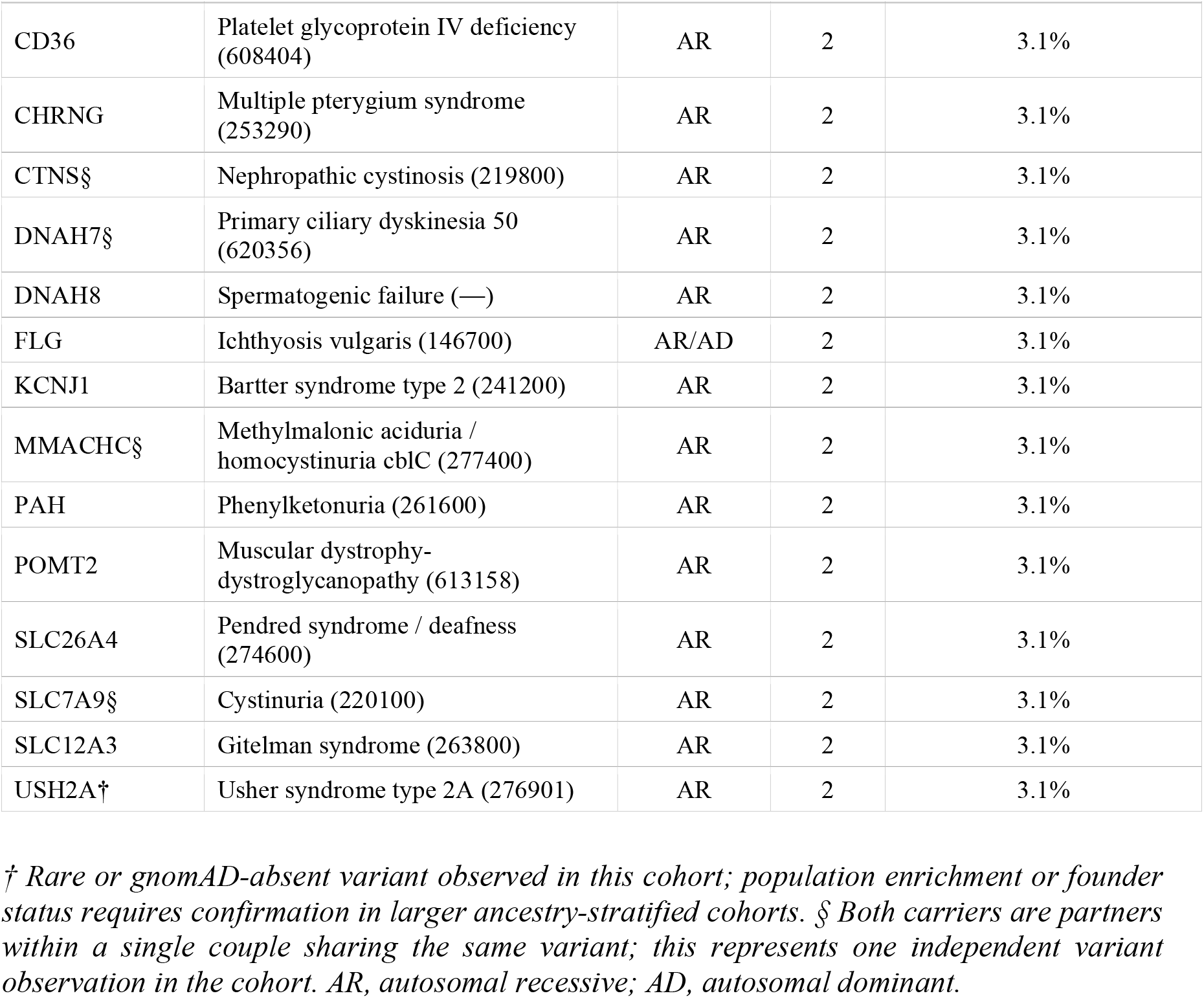
Genes with P/LP variants detected by WES in ≥2 of 65 WES-screened participants.

### SMA Carrier Screening

Among 179 IMC patients who underwent SMA carrier screening, four carriers were identified (2.2%; 95% CI 0.9–5.6%): two regular carriers (1+0 genotype; identified through SMA-only testing outside the WES cohort) and two silent carriers (2+0 genotype; Participants SMA-SC1 and SMA-SC2, both members of the WES cohort of 65).

Silent carriers harbour two SMN1 copies in cis on one chromosome and zero on the other. Standard copy-number testing would report a total copy number of two and classify these individuals as non-carriers; the 2+0 configuration was revealed only through the CarrierMax assay’s simultaneous detection of the g.27134T>G intragenic SNP combined with the exon 7 deletion g.27706_27707delAT. Both silent carriers carry reproductive risk for SMA-affected offspring equivalent to regular carriers if their partner is also an SMA carrier. Partner testing in both cases confirmed non-carrier status in the respective partner, excluding reproductive SMA risk for those couples. The two regular carriers were identified in the broader SMA-tested population outside the WES cohort of 65.

### Fragile X Syndrome Screening

Among 155 IMC females who underwent FMR1 CGG repeat analysis, one participant was found to carry an intermediate CGG allele of exactly 45 repeats with a second allele of 30 repeats (0.6%; 95% CI 0.1–3.6%). The 45-repeat allele falls at the lower boundary of the intermediate range (45–54 CGG) as defined by ACMG guidelines. No premutation (55–200 CGG) or full mutation (>200 CGG) alleles were identified. This individual was counselled regarding the minimal but non-zero risk of repeat expansion in future generations, primarily through maternal transmission. She is not a member of the WES cohort of 65.

### DMD Carrier Screening

MLPA identified no DMD exonic deletions or duplications in 29 preconception females. Among the 15 who also underwent WES, no reportable DMD sequence variants were detected. For the remaining 14 individuals, MLPA-negative status excludes detectable exonic deletions or duplications but does not exclude DMD point variants or small indels.

### Couple-Level Concordance Analysis

Of the 19 couples in whom both partners completed WES-based carrier screening, four concordant at-risk couples were identified (21.1%; 95% CI 8.5–43.3%).

#### Couple C1

Both partners carry CTNS c.225+1G>A (P/LP; absent from gnomAD v4.1; ClinVar submissions: 1 pathogenic, 1 likely pathogenic), conferring 25% per-pregnancy risk for nephropathic cystinosis (MIM 219800). Both additionally carry SLC7A9 c.459C>A (p.Cys153Ter; LP), conferring an independent 25% per-pregnancy risk for cystinuria (MIM 220100). This couple faces elevated preconception risk for two independent AR conditions simultaneously. Post-test genetic counseling addressed both findings.

#### Couple C2

Both partners carry MMACHC c.331C>T (p.Arg111Ter; P; ClinVar ID 1424; criteria PVS1, PS4_Moderate, PM2_Supporting), conferring 25% per-pregnancy risk for methylmalonic aciduria and homocystinuria, cblC type (MIM 277400). A combined couple report documented a 1-in-4 reproductive risk. Both partners were counselled and referred for PGT-M.

#### Couple C3

Both partners carry DNAH7 c.5869C>T (p.Arg1957Ter; LP; absent from gnomAD v4.1; criteria PVS1, PM2_Supporting), conferring 25% per-pregnancy risk for primary ciliary dyskinesia type 50 (MIM 620356). The male partner additionally carries a hemizygous G6PD pathogenic variant, addressed separately in counseling. Both partners were referred for PGT-M.

#### Couple C11

Both partners of Couple C11 carry LP variants in POMT2 (NM_013382.7; MIM 613158), conferring a 25% per-pregnancy risk of compound heterozygous POMT2-related muscular dystrophy-dystroglycanopathy. Partner C11-A carries NM_013382.7:c.320C>T (p.Pro107Leu; LP; ClinVar ID 95543; rs398124264; gnomAD 0.003%; criteria PM2, PP3, PP2). Partner C11-B carries NM_013382.7:c.878T>C (p.Leu293Pro; LP; absent from gnomAD; not in ClinVar; criteria PM2, PP3, PP2). In addition, Partner C11-A carries CFTR c.1367T>C (p.Val456Ala; Pathogenic) and Partner C11-B carries CFTR c.2562_2563delinsGA (p.Val855Ile; VUS; ClinVar 647129); as the latter variant is classified VUS per laboratory ACMG/AMP criteria, this CFTR co-occurrence does not meet the study’s formal concordance definition but represents an additional clinical finding discussed during counseling. Both partners were counselled; PGT-M referral is under discussion given the POMT2 concordant finding and additional CFTR carrier status in one partner.

Among the remaining 15 couples, two included a silent SMA carrier (Participants SMA-SC1 and SMA-SC2) whose partners were confirmed non-carriers by SMA testing, excluding reproductive SMA risk in both cases. In one couple (Couple C4), each partner carries LP variants in different AR disease genes (ACADSB and SLC12A3/IGHMBP2/DTYMK respectively); cross-partner genotyping is ongoing. Two recurring pathogenic variants were identified across unrelated participants: C8B c.1282C>T (p.Arg428Ter; C8 deficiency type II, MIM 613789) and ARSA c.542T>G (p.Ile181Ser; metachromatic leukodystrophy, MIM 250100) were each found independently in two participants of different ethnic backgrounds, raising the possibility of regional enrichment and warranting evaluation in larger ancestry-stratified cohorts.

## Discussion

We present, to our knowledge, the first reported WES-based multi-platform preconception carrier screening clinical program from Uzbekistan and Central Asia. A PubMed and Google Scholar search (search terms: Central Asia OR Uzbekistan AND carrier screening OR preconception; conducted May 2026) identified no prior published clinical implementation studies of WES-based multi-platform PCS from this region. Several findings are of particular scientific and clinical significance.

### High Carrier Detection Rate

A carrier detection rate of 93.8% (61/65) is consistent with published figures for expanded carrier screening performed using full whole-exome sequencing data (~20,000 genes). The high detection rate reflects the analytical breadth of bioinformatics filtering applied to the entire exome followed by clinical geneticist interpretation rather than an elevated population-specific pathogenic variant burden per se. The mean of 2.5 reportable P/LP variants per carrier across 126 unique genes underscores the genetic heterogeneity observed when analyzing the full exome.

### CFTR Allelic Spectrum

CFTR was the most frequently implicated gene (5/65 participants, 7.7%). Notably, none of the five P or LP variants identified represents a classical CF-causing allele such as F508del, W1282X, or G542X, prevalent in European and many other populations. The variant c.3935A>G (p.Asp1312Gly; ClinVar 53857) was explicitly characterized in the laboratory report as associated with CFTR-related disorders — including congenital bilateral absence of the vas deferens (CBAVD) and bronchiectasis — rather than classic cystic fibrosis. A second variant, c.2856G>C (p.Met952Ile; ClinVar 53580), carries conflicting ClinVar classifications (9 Pathogenic, 10 Likely Pathogenic, 3 VUS submissions) and was classified Likely Pathogenic based on PM3_Strong evidence (seen in trans with CF alleles in CBAVD patients). The remaining three variants were classified P or LP on ACMG/AMP criteria. Cross-referencing against the CFTR2 database (cftr2.org) classified c.413_415dup (p.Leu138dup; ClinVar 53905) as a CF-causing allele associated with classical cystic fibrosis, c.3154T>G (p.Phe1052Val) as varying clinical consequence, and c.1367T>C (p.Val456Ala) as CF-causing; c.3935A>G and c.2856G>C are not listed in CFTR2 with established CF-causing or varying-clinical-consequence status and should be considered of uncertain CF-disease relevance pending further population-level evidence. One additional CFTR variant, c.2562_2563delinsGA (p.Val855Ile; ClinVar 647129), was identified in Partner C11-B and initially recorded as LP; re-review against the signed laboratory report confirmed laboratory classification as VUS (ClinVar: 1 Likely Benign, 4 VUS; criteria PM2, PP2 only), and this variant has been reclassified accordingly and excluded from the carrier count. The overall CFTR allelic spectrum — dominated by CFTR-related disorder variants rather than classic CF alleles — may reflect the population-specific variant landscape of Central Asian populations and warrants further study.

### GJB2 and Non-Syndromic Hearing Loss

GJB2 was identified as the second most frequently implicated carrier gene (4/65, 6.2%), with all four variants being distinct — c.35del (35delG), c.−23+1G>A, c.101T>C, and c.95G>A. The 35delG variant (c.35del) is the most common GJB2 pathogenic variant worldwide and accounts for the majority of GJB2-associated hearing loss in European and Middle Eastern populations; its detection in this cohort reflects its pan-ethnic frequency. The high overall GJB2 carrier frequency is consistent with published estimates of 1–2% carrier frequency per variant globally, and aggregate GJB2 carrier rates of 2–6% in diverse populations. GJB2 carrier screening is therefore well justified in this population and should be retained as a high-priority gene in any regionally adapted screening program.

### Rare Recurrent Variants and Population-Genomic Considerations

Several rare or gnomAD-absent variants of regional interest were observed. USH2A c.13339A>G was identified in two unrelated participants; CTNS c.225+1G>A was found in both members of one couple; and PLEKHG2 c.610C>T and LIPH c.742C>A were each identified in one participant. These findings are hypothesis-generating rather than definitive: with n=65, mixed ethnicity, and no haplotype analysis, formal founder or population-enriched allele designation is premature. Confirmation in larger ancestry-stratified Central Asian cohorts with haplotype analysis is required. Nevertheless, the recurrent USH2A finding and the shared couple-level CTNS finding support prioritization of Central Asian population genomic reference data.

### Silent SMA Carriers and the Limitation of Standard Copy-Number Testing

Two of the four SMA carriers in this cohort were silent carriers (2+0 genotype), detected only through the intragenic SNP analysis incorporated in the CarrierMax assay. Both would have been classified as non-carriers by any standard copy-number-only testing approach reporting a total of two SMN1 copies. This finding illustrates a critical limitation of qPCR-based or SNP-unaware copy-number methods for SMA carrier screening in populations where the 2+0 haplotype may have appreciable frequency. The proportion of silent carriers among total SMA carriers in our cohort (50%) is notably higher than the ~8% reported for Ashkenazi Jewish populations and is consistent with higher proportions observed in certain Asian populations where the SMN1 duplication allele is more prevalent (Luo et al. 2014; Sugarman et al. 2012). Programs in Central Asia and similar regions should use assay systems capable of detecting the 2+0 genotype.

### Concordant At-Risk Couple Rate and Consanguinity

Although based on a small number of couples, the observed concordant at-risk couple rate of 21.1% (4/19; 95% CI 8.5–43.3%) is higher than the 2–4% reported in outbred IVF and general population cohorts (Capalbo et al. 2021; Tong et al. 2022). This elevated rate, while based on a small number of couples and subject to wide confidence intervals, is biologically plausible given Uzbekistan’s consanguinity prevalence. The finding that three of the four concordant couples involved variants absent from gnomAD (CTNS c.225+1G>A, DNAH7 c.5869C>T, and POMT2 c.878T>C) further supports a role for founder effects in driving couple-level concordance in this setting. All four at-risk couples were referred for counseling and two for PGT-M, representing direct clinical outcomes from a WES-based multi-platform PCS program not previously reported from this region. PGT-M is available at several accredited IVF clinics within Uzbekistan; both referred couples were directed through established local referral pathways. The availability of in-country PGT-M represents an important component of a comprehensive reproductive genetics service for Uzbek families identified at reproductive risk.

### Zero DMD Carrier Rate

No DMD exonic deletions or duplications were identified in 29 preconception females (0%; 95% CI 0.0–11.7%). The upper confidence bound of 11.7% reflects the small sample size; this cohort is insufficient to draw conclusions about population-specific DMD carrier frequency. Published estimates for DMD MLPA-detectable carrier frequency in females from diverse populations are approximately 1/3,000–1/5,000 live births (affected male prevalence) × 3 (carrier frequency approximation), or ~0.06–0.10%. A negative result in 29 individuals is consistent with these estimates but does not exclude DMD carrier prevalence at expected levels. The dual-platform coverage provided by WES (for point mutations and small indels) combined with MLPA (for large deletions and duplications) provides the most comprehensive available approach for DMD carrier detection.

### FMR1 Intermediate Allele

The identification of a single FMR1 intermediate allele (45 CGG repeats) in 155 females (0.6%) is consistent with published intermediate allele frequencies of approximately 0.8–1.1% in various populations, including 1/130 (0.77%) in Chinese women and 1.08% in Thai blood donors (Gao et al. 2020; Hnoonual et al. 2024; Nolin et al. 2019). At exactly 45 repeats — the lower boundary of the intermediate range — the risk of expansion to a premutation in a single generation is very low but non-zero, particularly through maternal transmission. No premutation or full mutation carriers were identified, consistent with the expected prevalence of approximately 1/259 females for premutation carriers in the general population (Hantash et al. 2011).

### Limitations

Several limitations of this study warrant acknowledgment. First, the cohort size of 65 WES-screened individuals is modest; estimates of gene-specific carrier frequencies and particularly of concordant couple rates are subject to wide confidence intervals. Second, the menu-based model means that not all participants received all assays, resulting in different denominators across platforms. Third, gnomAD v4.1 provides limited reference data for Central Asian populations; allele frequencies for several variants reported here are based on South Asian or East Asian proxy populations or are absent from the database entirely, limiting pathogenicity assessment. Fourth, a subset of variants — particularly those classified as LP on limited criteria (PM2 alone, or PM2+PP3) — may be reclassified as VUS on future evidence. Fifth, the cohort’s ethnic diversity, while representative of the clinical population at IMC Genomics Tashkent, limits direct extrapolation of population-specific carrier frequencies to the Uzbek population as a whole. The current program does not include dedicated haemoglobinopathy screening (e.g., HPLC or capillary electrophoresis); α-thalassemia in particular is poorly detected by WES due to its large-deletion mechanism, and dedicated haemoglobinopathy testing is a planned program expansion given the expected relevance in Central Asian populations. No MEFV P/LP variants were identified in this cohort; given expected enrichment of familial Mediterranean fever variants in Turkic populations, this likely reflects the small sample size and warrants prospective evaluation. Consanguinity status was not systematically recorded for all participants; prospective pedigree data collection would strengthen interpretation of couple-level concordance findings.

## Conclusion

This first reported WES-based multi-platform preconception carrier screening program in Uzbekistan demonstrates that expanded carrier screening combined with targeted assays for SMA, FMR1, and DMD is feasible and clinically productive in a Central Asian setting. The high carrier detection rate, the identification of hypothesis-generating rare or gnomAD-absent variants, the 21.1% concordant at-risk couple rate, and the detection of two SMA silent carriers who would have been missed by standard copy-number testing all underscore the clinical value of expanded, technologically comprehensive approaches in a population with high consanguinity prevalence and a largely uncharacterized variant landscape. These findings provide an evidence base for establishing structured preconception carrier screening as a component of reproductive healthcare in Uzbekistan and the broader Central Asian region, and highlight the need for population-specific genomic reference data for this understudied population.

## Statements and Declarations

### Ethics Approval and Consent to Participate

This study was conducted in accordance with the Declaration of Helsinki. Retrospective analysis of de-identified clinical data was approved by the IMC Genomics Ethics Review Board (approval no. 2025-04, dated 3 April 2025), an institutional review body operating independently of the clinical laboratory under the governance framework of Israeli Medical Center, Tashkent. All participants provided written informed consent at the time of sample collection for genetic testing and for the anonymized use of their clinical and genetic data for research and publication purposes, including publication of de-identified variant-level genetic data in supplementary tables. No identifiable personal information is included in this manuscript or its supplementary materials; study identifiers (e.g., C1-A, P001) are internal codes not linked to any external registry.

### Author Contributions

A. Kullyev: conceptualization, study design, clinical interpretation, manuscript preparation. S. Avdeichik: clinical data collection, genetic counseling, result interpretation, clinical interpretation. A. Akimenkova: laboratory analysis, variant classification. A. Kartuesov: laboratory analysis. O. Kardymon: bioinformatics analysis, variant annotation. Y. Goikhman: project supervision, clinical oversight, critical manuscript revision. All authors reviewed and approved the final manuscript.

## Acknowledgements

The authors thank all participants of the IMC Genomics preconception carrier screening program for consenting to the anonymized use of their clinical and genetic data for research and publication purposes, including publication of de-identified variant-level data.

## Data Availability

De-identified variant-level data supporting the findings of this study are presented in Supplementary Tables 1 and 2. Patient-level clinical data are not publicly available due to privacy and ethical constraints. Novel and rare variants of regional interest identified in this study, including USH2A c.13339A>G, CTNS c.225+1G>A, DNAH7 c.5869C>T, and PLEKHG2 c.610C>T, will be submitted to the NCBI ClinVar database (https://www.ncbi.nlm.nih.gov/clinvar/) prior to publication; ClinVar submission accession numbers will be provided in the final published version.

