## Supplementary Tables 1 and 2 for "Whole-exome-based preconception carrier screening in Uzbekistan with targeted SMA, FMR1, and DMD assays: the first reported clinical program"

**Supplementary Table 1: Complete inventory of pathogenic and likely pathogenic variants identified by whole-exome sequencing (WES cohort, n=65). All variants satisfied laboratory-validated quality metrics for high-confidence NGS reporting (mean coverage ~160×, ≥93% of targeted bases at ≥30×).**

| Study ID | Gene | MIM | HGVS / Variant | Classification | Run | ClinVar ID |
|---|---|---|---|---|---|---|
| C1-A | CTNS | 219800 | c.225+1G>A (splice) | P/LP | 1 | — |
| C1-A | SLC7A9 | 220100 | c.459C>A (p.Cys153Ter) | LP | 1 | — |
| C1-A | PLEKHG2† | 616763 | c.610C>T (p.Arg204Trp) | LP | 1 | — |
| C1-B | CTNS | 219800 | c.225+1G>A (splice) | P/LP | 1 | — |
| C1-B | SLC7A9 | 220100 | c.459C>A (p.Cys153Ter) | LP | 1 | — |
| P001 | EXOC3L2 | — | c.22G>T (p.Gly8Ter) | P | 3 | — |
| P001 | MYH3 | — | c.-9+1G>A (splice) | LP | 3 | — |
| P001 | FREM1 | — | c.3224dup (p.Asn1075fs) | LP | 3 | — |
| C5-A | RPGRIP1L | 611560 | c.3432+1G>A (intron 23) | LP | 4 | rs2150974573 |
| C5-A | TNNT1 | 605355 | c.750+1G>C (intron 12) | LP | 4 | rs111998831 |
| C5-B | GJB2 | 220290 | c.101T>C (p.Met34Thr) | LP | 4 | rs35887622 |
| C6-A | GJB2 | 220290 | c.-23+1G>A | P | 4 | 17029 |
| C6-B | LAMB2 | 614199 | c.4574-1G>C (splice) | LP | 4 | — |
| C6-B | MCCC2 | 210210 | c.1015G>A (p.Val339Met) | P | 4 | 203805 |
| C6-B | SGSH | 252900 | c.697C>T (p.Arg233Ter) | P | 4 | 370732 |
| C7-A | LIPA | 278000 | c.932G>A (p.Gly311Glu) | P | 4 | — |
| C7-B | FLG | 146700 | c.7339C>T (p.Arg2447Ter) | P | 4 | 50932 |
| C7-B | CFAP57 | 620917 | c.2554C>T (p.Gln852Ter) | LP | 4 | — |
| C8-A | — | — | NO FINDINGS (confirmed adequate QC) | — | 5 | — |
| C8-B | CFTR | 219700 | c.3935A>G (p.Asp1312Gly)* | P | 5 | 53857 |
| C8-B | SLC5A2 | 233100 | c.885+5G>A | LP | 5 | 632249 |
| P002 | CNGA3 | 216900 | NM_001298.3:c.485A>T (p.Asp162Val) | P | 6 | — |
| P002 | DNAH9 | 612444 | NM_001372.4:c.8839C>T (p.Arg2947Ter) | LP | 6 | — |
| P002 | PAH | 261600 | c.355C>T (p.Pro119Ser) | LP | 6 | — |
| P002 | TAT | 276600 | c.412G>A (p.Val138Ile) | LP | 6 | — |
| C9-A | HOGA1 | 613616 | NM_138413.4:c.700+5G>T (splice, intron 8) | P | 6 | rs185803104 |
| C9-B | NUP93 | 600800 | c.2137-18G>A (splice) | LP | 6 | — |
| C9-B | CLCN1 | 255700 | c.1936A>G (p.Met646Val) | LP | 6 | — |
| P003 | CYP21A2 | 201910 | c.518T>A (p.Ile173Asn) | P | 6 | — |

| Study ID | Gene | MIM | HGVS / Variant | Classification | Run | ClinVar ID |
|---|---|---|---|---|---|---|
| C10-A | MYO1E | 614868 | c.2706del (p.Phe903fs) | LP | 7 | — |
| C10-A | CHST6 | 217800 | c.382G>A (p.Ala128Thr) | LP | 7 | 3581262 |
| C10-B | KLHL40 | 615340 | c.176G>C (p.Arg59Pro) | LP | 7 | 807621 |
| C10-B | ACY1 | 609924 | c.1063-1G>A (splice) | LP | 7 | — |
| C10-B | DNHD1 | 619712 | c.5785C>T (p.Arg1929Ter) | LP | 7 | — |
| C10-B | PYGL | 232700 | c.1195C>T (p.Arg399Ter) | P | 7 | 21327 |
| C11-A | CFTR | 219700 | NM_000492.4:c.1367T>C (p.Val456Ala) | P | 9 | rs193922500 |
| C11-A | KCNJ1 | 241200 | c.601C>T | LP | 9 | — |
| C11-A | POMT2 | 613158 | NM_013382.7:c.320C>T (p.Pro107Leu) | LP | 9 | — |
| C11-B | POMT2 | 613158 | c.878T>C (p.Leu293Pro) | LP | 9 | — |
| C11-B | PTPRQ | 613391 | NM_001145026.2:c.1918_1921dup (p.Ala641Glyfs*13) | LP | 9 | 35821 |
| C11-B | AMPD1 | 615511 | c.468G>T (p.Gln156His) | LP | 9 | — |
| C11-B | CFTR | 219700 | NM_000492.4:c.2562_2563delinsGA (p.Val855Ile) | VUS* | 9 | 647129 |
| P004 | ABCA4 | 248200 | c.4919G>A (p.Arg1640Gln) | LP | 9 | 61751403 |
| P004 | FLG | 146700 | c.12064A>T (p.Lys4022Ter) | LP | 9 | 146466242 |
| P004 | SCN9A | 243000 | c.3482G>A (p.Trp1161Ter) | LP | 9 | 759003928 |
| P004 | CENPJ | 608393 | c.2372_2373insAA + c.2373_2374insTAA | LP | 9 | multiple |
| P005 | DNAH8 | — | NM_001206927.2:c.3483_3489del (p.Thr1162LysfsTer7) | P | 10 | — |
| P005 | GLDN | 617870 | NM_181789.4:c.542-1G>C (splice acceptor) | LP | 10 | — |
| P006 | DUOX2 | 274900 | NM_001363711.2:c.477del (p.Glu160ArgfsTer16) | P | 10 | — |
| P006 | SOHLH1 | — | c.947-1G>A (splice) | LP | 10 | — |
| P007 | ALDOB | 229600 | c.524C>A | P | 10 | — |
| P007 | PMFBP1 | — | c.2725C>T | LP | 10 | — |
| P007 | SPEF2 | — | c.3337C>T | LP | 10 | — |
| P008 | EXOSC3 | 614345 | c.238G>T | P | 10 | — |
| P008 | LIPH† | 604379 | c.742C>A (p.Leu248Met) | P | 10 | — |
| P008 | ATP7B | 277900 | c.2621C>T | P | 10 | — |
| P008 | CHRNG | 253290 | c.1249+2T>C | LP | 10 | — |
| P009 | USH2A† | 276901 | NM_206933.4:c.13339A>G (p.Met4447Val) | P | 10 | — |

| Study ID | Gene | MIM | HGVS / Variant | Classification | Run | ClinVar ID |
|---|---|---|---|---|---|---|
| P009 | STIL | 608647 | NM_001048166.1:c.3838C>T (p.Arg1280Cys) | LP | 10 | — |
| P010 | ATP7B | 277900 | NM_000053.4:c.3316G>A (p.Val1106Ile) | P | 10 | — |
| P010 | DNAH8 | — | NM_001206927.2:c.3904C>T (p.Arg1302Ter) | LP | 10 | — |
| P011 | OTOF | 601071 | NM_194248.3:c.583+1G>T | LP | 10 | — |
| P011 | GEMIN5 | 619333 | NM_015465.5:c.2196dup (p.Arg733fs) | LP | 10 | — |
| P011 | FIG4 | 611228 | NM_014845.6:c.1141C>T (p.Arg381Ter) | P | 10 | 217228 |
| P011 | PHYH | 266500 | NM_006214.4:c.42_60dup (p.Ser21fs) | P | 10 | 1454975 |
| P012 | WNT10A | 189550 | c.682T>A | LP | 10 | — |
| P013 | SLC26A2 | 222600 | NM_000112.4:c.-26+2T>C (splice donor) | P | 10 | — |
| P013 | GALNS | 253000 | NM_000512.5:c.374C>T (p.Pro125Leu) | LP | 10 | — |
| P013 | CHRNG | 253290 | NM_005199.5:c.1249+2T>C (splice donor) | P | 10 | — |
| P013 | LOXHD1 | 614934 | NM_001384474.1:c.4212+1G>A (splice donor) | LP | 10 | — |
| P013 | ALDOB | 229600 | c.524C>A | LP | 10 | — |
| P013 | ABHD12 | 612674 | NM_001042472.3:c.755C>T (p.Ala252Val) | LP | 10 | — |
| P014 | CEP295 | — | c.5045dup | LP | 10 | — |
| P014 | ABCA1 | 205400 | c.2804A>G | LP | 10 | — |
| P014 | PLA2G6 | 256600 | c.1547_1548dup | LP | 10 | — |
| P014 | CFAP298 | 615408 | c.292C>T | LP | 10 | — |
| P015 | CD36 | 608404 | c.610-2A>G (splice) | P | 10 | — |
| C2-A | MMACHC | 277400 | c.331C>T (p.Arg111Ter) | P | 11 | 1424 |
| C2-A | CNGB3 | 262300 | c.1077C>G (p.Tyr359Ter) | LP | 11 | — |
| C2-A | MSTO1 | — | c.268C>T (p.Gln90Ter) | LP | 11 | — |
| C2-B | MMACHC | 277400 | c.331C>T (p.Arg111Ter) | P | 11 | 1424 |
| C2-B | SLC34A3 | 241530 | c.1481del | LP | 11 | — |
| C2-B | F7 | 227500 | c.149C>G | LP | 11 | — |
| C2-B | GRM6 | — | c.1544C>A | LP | 11 | — |
| C2-B | FCSK | — | c.1989-2A>T (splice) | LP | 11 | — |
| C3-A | OCA2 | 203200 | c.1255C>T (p.Arg419Trp)‡ | P | 11 | — |
| C3-A | DNAH7 | 620356 | c.5869C>T (p.Arg1957Ter) | LP | 11 | — |
| C3-A | SLX4 | 613951 | c.4394_4404del | LP | 11 | — |

| Study ID | Gene | MIM | HGVS / Variant | Classification | Run | ClinVar ID |
|---|---|---|---|---|---|---|
| C3-A | ALDH4A1 | 239510 | c.802del | LP | 11 | — |
| C3-B | DNAH7 | 620356 | c.5869C>T (p.Arg1957Ter) | LP | 11 | — |
| C3-B | OAT | 258870 | c.1250C>T | LP | 11 | — |
| C3-B | CBLIF | 261000 | c.137C>T | LP | 11 | — |
| C3-B | RAG1 | 601457 | c.1520G>A | LP | 11 | — |
| C12-A | PEX6 | 614862 | c.1802G>A | P | 11 | — |
| C12-A | SLC25A13 | 605814 | c.1078C>T | P | 11 | — |
| C12-A | ABCA4 | 248200 | c.3481C>T | P | 11 | — |
| C12-A | SLC26A4 | 274600 | c.772C>T | LP | 11 | — |
| C12-A | ILDR1 | 614035 | c.772C>T | LP | 11 | — |
| C12-B | CFTR | 219700 | NM_000492.4:c.2856G>C (p.Met952Ile) | LP | 11 | 53580‡ |
| C12-B | CYP1B1 | 231300 | c.182G>A | LP | 11 | — |
| C12-B | KCNJ1 | 241200 | c.601C>T | LP | 11 | — |
| C13-A | TRAIP | 616777 | c.461del (p.Glu154GlyfsTer10) | LP | 12 | — |
| C13-B | MPZL2 | 618145 | c.72del | LP | 12 | — |
| C13-B | CSTA | 607936 | c.256C>T (p.Gln86Ter) | LP | 12 | — |
| C13-B | SC5D | 607330 | c.86G>A (p.Arg29Gln) | LP | 12 | — |
| P027 | ACADVL | 201475 | c.829_831del | LP | 12 | — |
| P027 | C6 | 612446 | c.2381+2T>C (splice) | LP | 12 | — |
| P027 | BLTP1 | 617822 | c.10022del (p.Gly3341GlufsTer26) | LP | 12 | — |
| C14-A | ISG15 | 616126 | c.3+1G>A (splice) | LP | 12 | — |
| C14-B | — | — | NO FINDINGS (confirmed adequate QC) | — | 12 | — |
| P016 | SLC12A3 | 263800 | NM_001126108.2:c.1924C>T (p.Arg642Cys) | LP | 12 | — |
| C4-A | ACADSB | 600301 | c.923G>A (p.Cys308Tyr) | LP | 12 | — |
| C4-B | SLC12A3 | 263800 | c.1456G>A | P | 12 | — |
| C4-B | IGHMBP2 | 604320 | c.2611+1G>A (splice) | P | 12 | — |
| C4-B | DTYMK | 617127 | c.20_29del | LP | 12 | — |
| C15-A | ASL | 207900 | c.1135C>G (p.Arg379Gly) | LP | 12 | — |
| C15-A | CD36 | 608404 | c.1156C>T (p.Arg386Trp) | LP | 12 | — |

| Study ID | Gene | MIM | HGVS / Variant | Classification | Run | ClinVar ID |
|---|---|---|---|---|---|---|
| C15-B | — | — | NO FINDINGS (confirmed adequate QC) | — | 12 | — |
| P017 | GALC | 245200 | c.136G>T (p.Asp46Tyr) | P | 12 | — |
| P018 | DNHD1 | 619712 | c.8782C>T (p.Arg2928Ter) | LP | 12 | — |
| P019 | USH2A† | 276901 | c.13339A>G (p.Met4447Val) | P | 12 | — |
| P020 | AAAS | 231550 | c.787T>C (p.Ser263Pro) | P | 12 | — |
| P020 | SLC26A4 | 274600 | c.1226G>A (p.Arg409His) | P | 12 | — |
| P021 | CFTR | 219700 | c.3154T>G (p.Phe1052Val) | P | 12 | — |
| P022 | C8B | 613789 | c.1282C>T (p.Arg428Ter) | P | 12 | 41286844 |
| P022 | CFTR | 219700 | c.413_415dup (p.Leu138dup) | P | 12 | — |
| P022 | ARSA | 250100 | c.542T>G (p.Ile181Ser) | P | 12 | — |
| P022 | IL11RA | 614732 | c.1-1G>T (splice) | LP | 12 | — |
| P023 | DHTKD1 | 614265 | c.1382del (p.Thr461AsnfsTer12) | LP | 13 | — |
| P023 | B3GALNT2 | — | c.979G>A (p.Asp327Asn) | LP | 13 | — |
| P023 | DNAH10 | — | c.5736C>G (p.Tyr1912Ter) | LP | 13 | — |
| P024 | C8B | 613789 | c.1282C>T (p.Arg428Ter) | P | 14 | 41286844 |
| P024 | GJB2 | 220290 | c.95G>A (p.Arg32His) | LP | 14 | rs111033190 |
| P025 | PYGM | 232600 | c.682del (p.Asp228IlefsTer67) | P | 14 | — |
| P025 | OTOG | 614945 | c.4657G>T (p.Gly1553Ter) | LP | 14 | — |
| P025 | SERPINC1 | 613118 | c.89T>A (p.Val30Glu) | LP | 14 | — |
| P025 | DYNC2H1 | 613091 | c.6035C>T | LP | 14 | — |
| C16-A | KIAA0586 | 616490 | NM_001329943.3:c.392del (p.Arg131LysfsTer4) | P | 14 | 204593 |
| C16-B | AP4B1 | 614066 | c.1160_1161del (p.Thr387ArgfsTer30) | LP | 14 | — |
| C16-B | GNRHR | 146110 | c.[317A>G;785G>A] (cis complex)§ | LP | 14 | — |
| P026 | — | — | NO FINDINGS (confirmed adequate QC) | — | 14 | — |
| C17-A | WEE2 | 617996 | c.224_227del (p.Glu75ValfsTer6) | LP | 14 | — |
| C17-A | DNAH5 | 608644 | c.12850dup (p.Tyr4284LeufsTer14) | LP | 14 | — |
| C17-A | C19orf12 | 614297 | c.172G>A (p.Gly58Arg) | LP | 14 | — |
| C17-A | ARSA | 250100 | c.542T>G (p.Ile181Ser) | LP | 14 | — |
| C17-A | GBE1 | 232500 | c.986A>G (p.Tyr329Cys) | LP | 14 | — |
| C17-B | FBXO7 | 260300 | c.1492C>T (p.Arg498Ter) | P | 14 | — |
| C17-B | MLH3 | 614385 | c.1755del (p.Glu586AsnfsTer24) | LP | 14 | ¶ |

| Study ID | Gene | MIM | HGVS / Variant | Classification | Run | ClinVar ID |
|---|---|---|---|---|---|---|
| C18-A | DNAI1 | 244400 | c.48+2dup (splice) | LP | 14 | — |
| C18-A | WFDC2 | 620984 | c.145T>C (p.Cys49Arg) | LP | 14 | — |
| C18-B | HFE | 235200 | c.845G>A (p.Cys282Tyr) | P | 14 | rs1800562 |
| C18-B | C2 | 217000 | c.841_849+19del | LP | 14 | — |
| C19-A | GJB2 | 220290 | c.35del (p.Gly12ValfsTer2) [35delG] | P | 14 | rs80338940 |
| C19-A | EIF2AK4 | 234810 | c.4462C>T (p.Gln1488Ter) | P | 14 | — |
| C19-A | PAH | 261600 | c.165T>G (p.Phe55Leu) | LP | 14 | — |
| C19-B | DNAAF4 | 615482 | c.733C>T (p.Arg245Ter) | P | 14 | — |
| C19-B | TTLL5 | 615860 | c.1627G>A (p.Glu543Lys) | LP | 14 | — |

*Abbreviations: P, Pathogenic; LP, Likely Pathogenic; VUS, Variant of Uncertain Significance; AR, autosomal recessive. † Rare or gnomAD-absent variant; population enrichment or founder status requires confirmation in larger ancestry-stratified cohorts with haplotype analysis. ‡ Conflicting ClinVar interpretations; laboratory-assigned classification shown. OCA2 c.1255C>T (p.Arg419Trp; C3-A; ClinVar 194160): 6 Pathogenic, 1 Likely Pathogenic, 3 VUS submissions. CFTR c.2856G>C (p.Met952Ile; ClinVar 53580; C12-B): 9 Pathogenic, 10 Likely Pathogenic, 3 VUS submissions (counts as of report date). § Complex cis allele on single chromosome (GNRHR c.[317A>G;785G>A]; C16-B); counted as one carrier event. ¶ MLH3 c.1755del (C17-B) is not on the ACMG SF v3.3 list; disclosed clinically. VUS* C11-B CFTR c.2562_2563delinsGA (ClinVar 647129): classified VUS by laboratory ACMG/AMP assessment (1 Likely Benign, 4 VUS submissions; criteria PM2, PP2 only); listed for clinical completeness and excluded from carrier frequency calculations. POMT2 carriers C11-A and C11-B are partners within a single couple carrying different POMT2 variants (each representing an independent observation). CTNS, SLC7A9, MMACHC, and DNAH7 carriers listed as partners within a single couple share the same variant (each representing one independent variant observation). G6PD c.1003G>A (p.Ala335Thr; WHO Class II; G6PD Chatham) hemizygous in participant C3-B (affected male genotype, not carrier) is omitted from this table; see main text. BTD c.1270G>C (p.Asp424His; pseudodeficiency allele) in participant C10-A is omitted from this table. All listed variants are heterozygous unless otherwise indicated.*

### Supplementary Table 2: ACMG Secondary Findings and Related Disclosures

| Study ID | Gene | Variant | Condition (MIM) | Inheritance | ACMG SF v3.3 | Note |
|---|---|---|---|---|---|---|
| SMA-SC1 (C5-A) | TTR | c.424G>A (p.Val142Ile) | Hereditary transthyretin amyloidosis (105210) | AD | Yes | gnomAD AFR freq >1.7%; disclosed clinically |
| C10-A | PMS2 | c.251-1G>T | Lynch syndrome / colorectal cancer (614385) | AD | Yes | Disclosed |
| P021 | SOD1 | c.272A>C | ALS type 1 (105400) | AD | Yes | Disclosed |
| P024 | CHEK2 | c.470T>C (p.Ile157Thr) | Hereditary breast/ovarian cancer | AD | Yes | Disclosed |
| C5-B | MYH7 | c.968T>C (p.Ile323Thr) | Hypertrophic cardiomyopathy (192600) | AD | Yes | Disclosed |
| C17-B | MLH3 | c.1755del (p.Glu586AsnfsTer24) | Lynch-like colorectal cancer (614385) | AD/AR | No | Not on ACMG SF v3.3; disclosed clinically given Lynch association |
